# Exploring Antimicrobial Resistance and Genetic Profile as Mortality and Morbidity Indicators in Pediatric Cystic Fibrosis

**DOI:** 10.1101/2025.09.22.25336281

**Authors:** Viraja Teggihal, Anagha Teggihal, Nishkala Uday Rao

## Abstract

Cystic Fibrosis is a multisystem disease that follows an autosomal recessive inheritance pattern. Pulmonary involvement is the most common cause of morbidity in these patients. In this study, we investigate the impact of antimicrobial resistance and genetic profile through a retrospective cohort study in 176 children from 2017-2021 in Western Romania. As treatment for Cystic Fibrosis infections is usually with the same group of antibiotics, we aimed to study antibiotic resistance as a whole. We observed that aminoglycoside resistance is associated with 35% respiratory exacerbations. Out of the 43 patients with aminoglycoside resistance, 15 experienced respiratory exacerbations. Although penicillin resistance is associated with increased mortality, in our study, we found that mortality was lower in the penicillin resistant cohort (38%) when compared to the penicillin non resistant cohort (54%). Out of 115 patients that exhibited penicillin resistance, 44 died and out of 61 patients that did not exhibit penicillin resistance, 33 died. On genetic profiling, we found that the DF508/N1303K subtype was protective while the DF508/C524X subtype was associated with increased mortality. More studies are needed to understand if they can be used as indicators of mortality and morbidity in pediatric cystic fibrosis outcomes.

**Conflicts of Interest:** The authors declare that there are no conflicts of interest regarding the publication of this manuscript.

## Introduction

Cystic Fibrosis (CF) involves various organs in the body and causes systemic complications. Lung involvement is the leading cause of morbidity and mortality [1] and more frequent respiratory infections and exacerbations have demonstrated worse clinical outcomes [2]. Decreased chloride excretion and unrestricted sodium absorption in the airway lining epithelium lead to the formation of large amounts of thick mucus, causing small airway plugging, irreversible lung damage, and respiratory failure [3].

Although pulmonary complications cause the majority of mortality in these children, many children can present with abdominal manifestations related to gastrointestinal and pancreaticobiliary disease. While neonates usually present with meconium ileus or its complications, older children tend to present with distal intestinal obstruction syndrome or colonic stricture as a result of high doses of pancreatic enzyme replacement [4].

With increasing age, the incidence of gastroesophageal reflux disease (GERD) increases reaching its peak by adulthood. In CF, patients with worsening lung disease and uncontrolled GERD, Nissen fundoplication is reported not only to help slow the decline in lung function but also cause significant improvements in weight and a reduction in CF exacerbations [5] and consequently the number of hospital admissions.

Well-known factors associated with Poor outcomes in CF patients include declining FEV 1 [6], infection with Pseudomonas aeruginosa, multiple hospitalizations [7], pancreatic insufficiency [8], age at diagnosis [9], homozygosity [10], pulmonary hypertension [11], clubbing or crackles [12], low socioeconomic status [13] and malnutrition [14]. The emergence of antimicrobial resistance (AMR) further complicates clinical outcomes and increases mortality rates.

This study examines the overall burden of AMR in pediatric cystic fibrosis patients, focusing on its impact on clinical outcomes. By analyzing resistance patterns across variables such as sex, cystic fibrosis subtype (genetic profile), respiratory exacerbations, and mortality, we highlight the critical role of resistance in shaping patient prognosis. Our study also aims to determine whether resistance to a particular antibiotic could serve as a mortality and morbidity indicator. Since multiple organisms can be resistant to the same antibiotics, evaluating resistance in this broader context is essential. While the emphasis is on the association of resistance with outcomes, we also acknowledge the role of specific pathogens, as variations in resistance mechanisms can influence outcomes as evidenced by various studies.

## Materials and Methods

### Study Design

It is a retrospective cohort study at the University of Medicine and Pharmacy ‘Victor Babes’ Timi□oara, Romania.

### Data Collection

Data was collected from electronic medical records from 2017-2021. Patient data like age, sex, genetic profile, antibiogram, respiratory exacerbation, and death were collected.

Inclusion criteria included a patient with confirmed or highly suspected CF from 0-18 years of age.

### Analysis Methods

#### a. Statistical Methods

Descriptive and inferential statistical analysis was carried out with all parameters. Results were reported in numbers and percentages with a level of significance of 5%

#### b. Assumptions

1. The dependent variables must follow a normal distribution.
2. Samples should be randomly selected from the population.
3. Each case within the samples should be independent.

#### c. Data Analysis

Chi-square test (Fisher’s exact test)

#### d. P value

1. 0.01< P ≤ 0.05: moderately significant
2. P≤0.01: strongly significant
3. Any other value was considered non-significant

#### e. Softwares used

SPSS 15.0, R environment ver.3.2.2, Microsoft Word and Excel.

## Results

### Genetic CF Subtype and Mortality

In our analysis, the Heterozygous DF508/N1303K subtype showed no recorded deaths (0% mortality, p=0.003), indicating a significant association with lower mortality. In contrast, the Heterozygous DF508/C524X subtype had a 100% mortality rate, with all four patients deceased (p=0.03). (Table 1)

**Table 1.**

| Genetic subtype of CF | Death |  | Grand Total | Fisher's Exact Test p-value |
| --- | --- | --- | --- | --- |
|  | No | Yes |  |  |
| Delta F508 (unspecified) | 10(10.1%) | 11(11.1%) | 21(21.2%) | 0.49 |
| Homozygous DF 508 | 23(23.2%) | 23(23.2%) | 46(46.5%) | 0.63 |
| Heterozygous DF 508 | 4(4%) | (0%) | 4(4%) | 0.13 |
| Heterozygous non-DF508 | 1(1%) | 5(5.1%) | 6(6.1%) | 0.09 |
| Heterozygous DF508/N1303K | 11(11.1%) | (0%) | 11(11.1%) | **0.003\*** |
| Heterozygous DF508/G542 | 7(7.1%) | 7(7.1%) | 14(14.1%) | 0.78 |
| Heterozygous DF508/1677 | 1(1%) | 2(2%) | 3(3%) | 0.58 |
| Heterozygous DF 508/L88X | 1(1%) | (0%) | 1(1%) | 1.0 |
| Heterozygous DF 508/X allele not identified | 2(2%) | (0%) | 2(2%) | 1.0 |
| Heterozygous DF 508/6542X | 1(1%) | (0%) | 1(1%) | 1.0 |
| Heterozygous DF508/G126D | 4(4%) | (0%) | 4(4%) | 0.13 |
| Heterozygous DF508/C524X | (0%) | 4(4%) | 4(4%) | **0.03\*** |
| Heterozygous DF 508/2184 | 1(1%) | (0%) | 1(1%) | 1.0 |
| Heterozygous DF508/D1152H | 1(1%) | (0%) | 1(1%) | 1.0 |
| Complete Clinical Form of Cystic Fibrosis | 25(25.3%) | 20(20.2%) | 45(45.5%) | 1.0 |
| suspected | 3(3%) | (0%) | 3(3%) | 0.26 |
| Cystic Fibrosis Clinical Pulmonary Form | 3(3%) | (0%) | 3(3%) | 0.26 |
| Cystic Fibrosis<br>Clinical Pancreatic<br>Form | 1(1%) | (0%) | 1(1%) | 1.0 |
| Heterozygous<br>DF508/non-DF508 | (0%) | 2(2%) | 2(2%) | 0.19 |
| Heterozygous<br>Compound Non-Delta<br>F508(G542X/Poly<br>morphic Allele IV<br>S8-9T) | (0%) | 3(3%) | 3(3%) | 0.08 |
| **Grand Total** | 99(100<br>%) | 77(77.8%) | 176(177.8%) |  |

### Gender and Mortality

Mortality rates did not significantly differ between male (50.6%) and female (49.4%) patients (p = 0.53), suggesting that gender does not play a significant role in determining mortality outcomes (Table 2) (Graph 1)

**Table 2:** Relationship of gender with mortality.

| Gender↓/ Death→ | No | Yes | Grand Total |
| --- | --- | --- | --- |
| **Female** | 51(51.5%) | 36(46.8%) | 87(49.4%) |
| **Male** | 48(48.5%) | 41(53.2%) | 89(50.6%) |
| **Grand Total** | 99(100%) | 77(100%) | 176(100%) |

Graph 1 (p = 0.53)

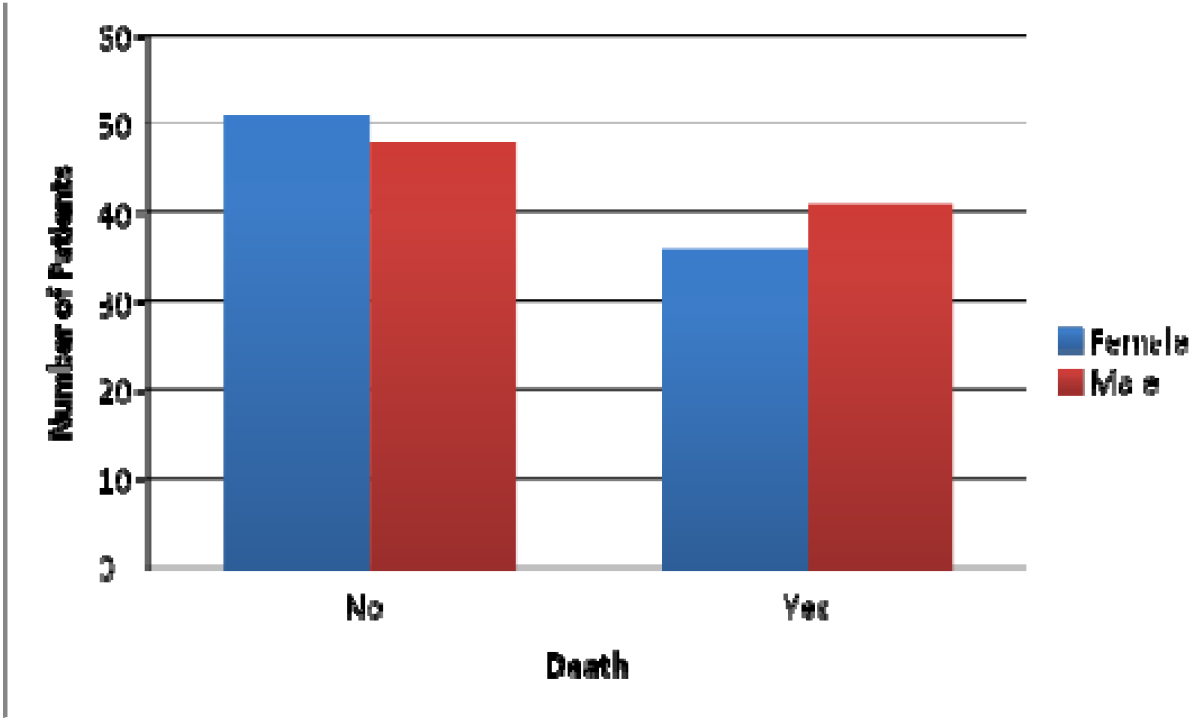

### Respiratory Exacerbations and Mortality

Patients experiencing respiratory exacerbations had a higher, though not statistically significant, mortality rate compared to those without exacerbations (p = 0.76). (Table 3) (Graph 2)

**Table 3:** Relationship of respiratory exacerbations with mortality.

| Respiratory<br>Exacerbation ↓/ Death → | No | Yes | Total |
| --- | --- | --- | --- |
| **No** | 79(79.8%) | 60(77.9%) | 139(79%) |
| **Yes** | 20(20.2%) | 17(22.1%) | 37(21%) |
| **Total** | 99(100%) | 77(100%) | 176(100%) |

Graph 2 (p = 0.76)

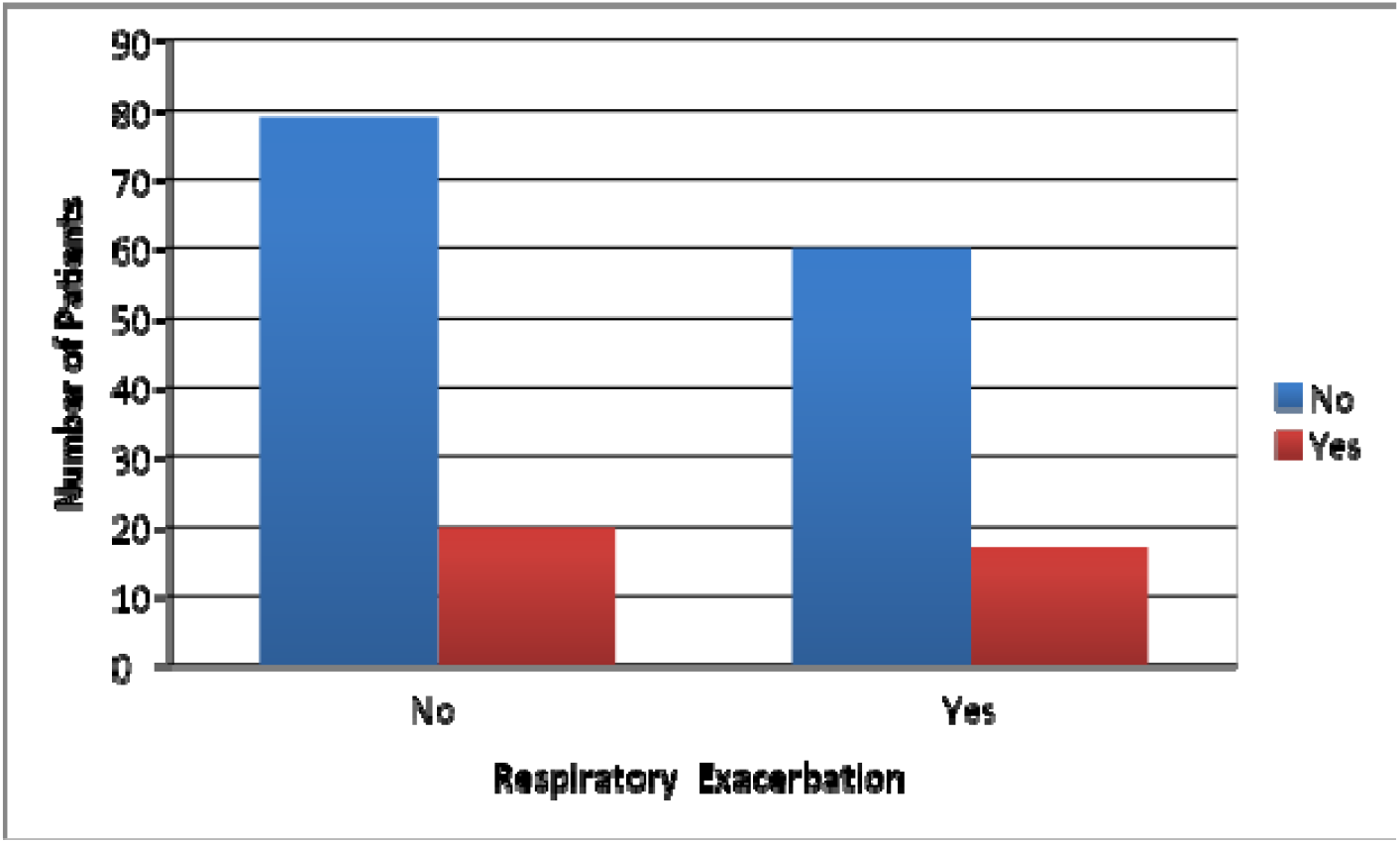

### Antimicrobial Resistance and Mortality

The analysis revealed that penicillin resistance was associated with a significantly higher mortality rate, with 54.1% of resistant patients dying compared to 38.26% of non-resistant patients (p-value 0.05). In comparison, mortality rates for other antibiotics, such as cephalosporins, fluoroquinolones, and antifungals, were also elevated but did not reach statistical significance. (Table 4)

**Table 4.**

| Antibiotics |  | Death |  | Grand Total | Fishers Exact test p-value |
| --- | --- | --- | --- | --- | --- |
|  |  | No | Yes |  |  |
| Cephalosporin | NO | 71(58.2%) | 51(41.8%) | 122(100%) | 0.51 |
|  | YES | 28(51.85%) | 26(48.15%) | 54(100%) |  |
| Fluoroquinolone | NO | 92(58.23%) | 66(41.77%) | 158(100%) | 0.13 |
|  | YES | 7(38.89%) | 11(61.11%) | 18(100%) |  |
| Carbapenem | NO | 93(56.71%) | 71(43.29%) | 164(100%) | 0.76 |
|  | YES | 6(50%) | 6(50%) | 12(100%) |  |
| Macrolide | NO | 61(52.14%) | 56(47.86%) | 117(100%) | 0.14 |
|  | YES | 38(64.41%) | 21(35.59%) | 59(100%) |  |
| Penicillin | NO | 28(45.9%) | 33(54.1%) | 61(100%) | 0.05* |
|  | YES | 71(61.74%) | 44(38.26%) | 115(100%) |  |
| Aminoglycoside | NO | 77(57.89%) | 56(42.11%) | 133(100%) | 0.48 |
|  | YES | 22(51.16%) | 21(48.84%) | 43(100%) |  |
| Lincosamide | NO | 66(54.55%) | 55(45.45%) | 121(100%) | 0.51 |
|  | YES | 33(60%) | 22(40%) | 55(100%) |  |
| Monobactam | NO | 99(56.9%) | 75(43.1%) | 174(100%) | 0.19 |
|  | YES | 0(0%) | 2(100%) | 2(100%) |  |
| Tetracycline | NO | 97(57.4%) | 72(42.6%) | 169(100%) | 0.24 |
|  | YES | 2(28.57%) | 5(71.43%) | 7(100%) |  |
| Colistin | NO | 96(56.47%) | 74(43.53%) | 170(100%) | 1 |
|  | YES | 3(50%) | 3(50%) | 6(100%) |  |
| Antifungal | NO | 96(58.18%) | 69(41.82%) | 165(100%) | 0.06 |
|  | YES | 3(27.27%) | 8(72.73%) | 11(100%) |  |

### Antimicrobial Resistance and Respiratory Exacerbation

The analysis showed that aminoglycoside resistance was significantly associated with an increased rate of respiratory exacerbations, with 34.88% of patients affected (p-value 0.01^*^). In contrast, resistance to other antibiotics, such as cephalosporins, showed lower rates of respiratory exacerbations, but these were not statistically significant. (Table 5)

**Table 5.**

| Antibiotics |  | Respiratory Exacerbation |  | Total | Fishers Exact test p-value |
| --- | --- | --- | --- | --- | --- |
|  |  | No | Yes |  |  |
| Cephalosporin | NO | 99(81.15%) | 23(18.85%) | 122(100%) | 0.3 |
|  | YES | 40(74.07%) | 14(25.93%) | 54(100%) |  |
| Fluoroquinolone | NO | 127(80.38%) | 31(19.62%) | 158(100%) | 0.21 |
|  | YES | 12(66.67%) | 6(33.33%) | 18(100%) |  |
| Carbapenem | NO | 132(80.49%) | 32(19.51%) | 164(100%) | 0.13 |
|  | YES | 7(58.33%) | 5(41.67%) | 12(100%) |  |
| Macrolide | NO | 89(76.07%) | 28(23.93%) | 117(100%) | 0.24 |
|  | YES | 50(84.75%) | 9(15.25%) | 59(100%) |  |
| Penicillin | NO | 46(75.41%) | 15(24.59%) | 61(100%) | 0.43 |
|  | YES | 93(80.87%) | 22(19.13%) | 115(100%) |  |
| Aminoglycoside | NO | 111(83.46%) | 22(16.54%) | 133(100%) | 0.01** |
|  | YES | 28(65.12%) | 15(34.88%) | 43(100%) |  |
| Lincosamide | NO | 93(76.86%) | 28(23.14%) | 121(100%) | 0.42 |
|  | YES | 46(83.64%) | 9(16.36%) | 55(100%) |  |
| Monobactam | NO | 137(78.74%) | 37(21.26%) | 174(100%) | 1 |
|  | YES | 2(100%) | (0%) | 2(100%) |  |
| Tetracycline | NO | 134(79.29%) | 35(20.71%) | 169(100%) | 0.63 |
|  | YES | 5(71.43%) | 2(28.57%) | 7(100%) |  |
| Colistin | NO | 133(78.24%) | 37(21.76%) | 170(100%) | 0.34 |
|  | YES | 6(100%) | (0%) | 6(100%) |  |
| Antifungal | NO | 131(79.39%) | 34(20.61%) | 165(100%) | 0.7 |
|  | YES | 8(72.73%) | 3(27.27%) | 11(100%) |  |

## Discussion

### Penicillin Resistance

Penicillin resistance in CF patients is usually exhibited by Methicillin Resistant Staphylococcus aureus (MRSA), Pseudomonas aeruginosa (PA), Acinetobacter baumannii, Haemophilus influenzae, Streptococcus pneumoniae, Enterobacteriaceae (ex: Klebsiella pneumoniae, Escherichia coli) and Burkholderia cepacia complex (Bcc).

Pulmonary infection by PA, Staphylococcus aureus, Aspergillus species, and mixed oral flora [15] are associated with significant inflammatory responses in young children with CF. MRSA infections are associated with more severe lung disease, increased hospitalization rates, and mortality rates [16]. These patients must be monitored regularly because over 25% of patients with CF in the United States have MRSA in their respiratory culture specimens [17].

A study conducted by Prevaes SM et al revealed that Infants with CF had an early dominance of Staphylococcus aureus, which then shifted to Streptococcus mitis while infants in the control group had Moraxella species and Hemophilus influenza consistently. Additionally, antibiotic use in CF infants increased harmful gram-negative bacteria like Burkholderia and reduced useful bacteria [18].

Various attempts have been made to overcome staphylococcal infections among CF patients. A review study concluded that antistaphylococcal antibiotic prophylaxis led to fewer children having isolates of Staphylococcus aureus when commenced early in infancy and continued up to the age of Its use however is debatable because of its uncertain clinical importance [19]. UK Cystic Fibrosis Trust supported this theory and recommended using antistaphylococcal antibiotic prophylaxis until the age of 3 [20].

Susceptibility testing of MRSA isolates from CF patients indicated that trimethoprim-sulfamethoxazole, tetracycline (either doxycycline or minocycline), and fusidic acid typically exhibited low rates of resistance in vitro. In contrast, there were high resistance rates (70-90%) to clindamycin and fluoroquinolones [21]. Novel antibiotics for MRSA include tedizolid, telavancin, dalbavancin, oritavancin, ceftobiprole, and daptomycin. None of these drugs have been approved for children so far [21].

A study by Godfrey AJ et al [22] found that high-dose piperacillin and tobramycin treatment led to the persistence of two PA serotypes, M and K, with increasing beta-lactam resistance. Serotype K showed low-level resistance with absent or altered penicillin-binding protein-3, while serotype M strains developed higher resistance, with reduced affinity for [14C] penicillin G. These changes suggested that resistance resulted from mutations during treatment. This further highlights the importance of the use of antibiotics only when necessary. Routine CF prophylaxis is not practised in the United States due to the lack of clear benefits.

In our study, penicillin resistance was linked to a notably high mortality rate of 54%, underscoring the need for further research into novel treatment approaches to reduce this burden.

### Aminoglycoside resistance

Mutations in the mexZ gene of PA lead to increased expression of mexY mRNA as well as increased resistance to amikacin [23]. For the first infection with PA, the preferred treatment is oral ciprofloxacin with inhaled tobramycin or inhaled colomycin usually but a trial by Early Pseudomonas Infection Control (EPIC) in 2014 on a group of children concluded that inhaled tobramycin irrespective of oral ciprofloxacin usage was effective[24]. The first-line treatment of choice for these infections is a combination therapy with beta-lactams and aminoglycosides, but imipenem or colistin with aminoglycosides has also proven to be effective [25, 26].

The rmtA gene is responsible for the broad-spectrum aminoglycoside resistance displayed by PA. Careful administration of aminoglycosides should be practised in clinical settings as this gene is a mobile genetic material and could transfer pan-aminoglycoside resistance across other species [27].

Aminoglycoside resistance is seen in a wide variety of organisms. Bcc, through efflux pumps, biofilms, and cell envelope modifications develop resistance [28]. Moreover, they survive the non-oxidative immune attacks from neutrophils while also combating against antimicrobial peptides that target the airway epithelial cells [29].

Achromobacter species [30, 31], especially Achromobacter xylosoxidans, is known to have a natural resistance to multiple drugs including broad-spectrum aminoglycosides and cephalosporins with an exception of ceftazidime [32, 33, 34, 35]. Acinetobacter baumannii is also known to have similar properties leading to poorer clinical outcomes [36].

The results of our study support the existing literature and show an increased respiratory exacerbation by 35% in aminoglycoside-resistant children.

### Gender

Women with CF are known to be colonised by pathogens much earlier than men leading to a decrease in life expectancy during these episodes or even otherwise despite consideration of co-morbidities related to CF [37]. A large-scale pediatric study by Rosenfeld et al also showed similar results [38]. While this is a commonly seen association, some studies have challenged it [39]. Similarly, in our study, mortality rates did not significantly differ across both the genders.

### Genetic Profile

A study [40] conducted on 17853 patients using the US Cystic Fibrosis Foundation National Registry revealed that the 11 most common genotypes, DeltaF508/R117H, DeltaF508/DeltaI507, DeltaF508/3849+10kbC-->T, and DeltaF508/2789+5G-->A had a significantly lower mortality rate than the genotype homozygous for DeltaF508. DeltaF508/R117H, DeltaF508/DeltaI507, DeltaF508/ 3849+10 kbC-->T, DeltaF508/2789+5G-->A, and DeltaF508/A455E have a milder clinical phenotype.

Patients with missense mutation R1066C in the second transmembrane domain of CFTR [41] and patients heterozygous for 3905insT [42] have been reported to have high morbidity and mortality comparable to patients homozygous for deltaF508.

Another study [43] investigating delta F508, R553X, and 3905insT reported significant differences in age at the onset of chronic PA colonization and Chrispin-Norman x-ray scores among the genetic subgroups. The age at P. aeruginosa colonization was noted to be 12 years in R553X homozygotes. R553X homozygotes had a two-stage disease progression: mild before colonization and severe after while R553X heterozygotes exhibited a milder clinical course. Delta F16 group showed more severe disease compared to Delta F508 homozygotes.

In our study, children with the heterozygous DF508/N1303K subtype showed no recorded deaths (0% mortality, p=0.003), indicating a significant association with lower mortality. In contrast, the heterozygous DF508/C524X subtype had a 100% mortality rate, with all four patients deceased (p=0.03). These findings suggest significant differences between subtypes and warrant further investigation.

## Conclusion

Our study is focused on the roles of antimicrobial resistance and various genetic subtypes as potential indicators in predicting the clinical outcomes in pediatric cystic fibrosis patients. While multiple studies have described the role of individual multidrug resistant organisms on poor outcomes, we aimed to explore resistance to antibiotics across all pathogens because treatment relies on the same class of antibiotics.

Resistance to penicillin and aminoglycoside antibiotics was significantly associated with increased mortality and respiratory exacerbations, respectively. Genetic profiling revealed the DF508/N1303K subtype being protective and the DF508/C524X subtype having a severe prognosis. However, more research is needed to better understand the early clinical indicators of pediatric cystic fibrosis patients.

## Data Availability

All data produced in the present work are contained in the manuscript

## Acknowledgements

We’d like to thank our professors, Dr. Monica Licker and Dr. Delia Muntean for their guidance and support in conducting this study.

